# Capturing India’s phenotypic diversity: Health insights from the GenomeIndia project

**DOI:** 10.64898/2026.04.01.26349926

**Authors:** Debasrija Mondal, Chandrika Bhattacharyya, Dolat S Shekhawat, Nayan G Tada, Tanuja Rajial, Arun Sree Parameswaran, Deepak Jena, Sudeshna Datta, Mamuni Swain, Sudarshan Jena, Adyasha Mishra, Soumendu Mahapatra, Shijulal Nelson Sathi, Mahabub Alam, Azad Ali, Parveena Choudhury, Poulomi Ghosh, Devashish Tripathi, Shobha Anilkumar, Divakar Ashwath, Mohana Chithimmaiah, Shafeeq K Shahul Hameed, Rajesh Gunasegaran, Neha Singh, G Mala, Tiyasha De, Shahrumi Reza, Ankit Mukherjee, Bhumika Prajapati, Bhagirath Dave, Silvia Yumnam, Kshetrimayum Vimi, Gurumayum Nikesh Sharma, Ajay Malik, Ranjan Jyoti Sarma, Andrew Vanlallawma, Doddaladka Krishnayya Samartha, S G Tejaswini, Paranthaman V Kavya, Sanjay Deshpande, GenomeIndia Consortium, Kuldeep Singh, Praveen Sharma, Sunil K Raghav, Punit Prasad, E V Soniya, Chaitanya Joshi, Madhvi Joshi, Nanaocha Sharma, Santosh Dixit, Mayurika Lahiri, L S Shashidhara, H Lalhruaitluanga, Lal Nundanga, Nachimuthu Senthil Kumar, Ganesan Venkatasubramanian, Naren P Rao, Venkataram Shivakumar, Mohd Ashraf Ganie, Imtiyaz Ahmad Wani, Arindam Maitra, Nidhan K Biswas, Bratati Kahali, Divya Tej Sowpati, Mohammad Faruq, Sridhar Sivasubbu, Vinod Scaria, Yadati Narahari, Kumarasamy Thangaraj, Vijayalakshmi Ravindranath, Prathima Arvind, Abdul Jaleel, Ganganath Jha, Suman Paine, Karthik Bharadwaj Tallapaka, Analabha Basu, Shweta Ramdas

## Abstract

**Background:** India represents 18% of the global population yet remains underrepresented in health research. Moreover, existing national surveys miss critical variation across its 4,600 ethnolinguistic groups. We present a comprehensive phenotypic characterisation of 81 populations from the GenomeIndia project.

**Methods:** We analysed 67 sociodemographic, anthropometric, and blood biochemistry variables from 17,777 individuals sampled across 81 ethnolinguistic populations from India, examining population-level variation, disease reporting fractions, and age- and sex-specific life-course trends.

**Findings:** Ethnolinguistic identity predicted health outcomes independently of administrative state, improving phenotypic variance explained by an average of 7·4%. 95% of participants had at least one abnormal biochemical or anthropometric marker, driven by low HDL (52·2%) and elevated triglycerides (43·6%). Metabolic risk, however, was highly stratified: adjusted prevalence for low HDL ranged four-fold across ancestry groups from 17·2% to 67·7%. We also identified an “awareness gap”; only 17·6% of people with hypertension and 2·2% of people with dyslipidemia were aware of their condition. This awareness gap was higher in tribal populations, in which women did not show the higher HDL levels typically seen compared to men, pointing to distinct metabolic profiles and healthcare access barriers across India.

**Interpretation:** The Indian phenotypic landscape is highly structured along ethnolinguistic lines, where ancestry and environment both influence risk. The high systemic burden of abnormalities necessitates population-specific reference intervals. GenomeIndia provides a foundational map for precision public health, shifting the focus from state-level averages to population-specific risk profiles.

## Introduction

Population-based epidemiological datasets are fundamental to studying health trends and disease patterns that inform public health interventions and policy. Such datasets across the world, such as the UK Biobank^1^, the China Kadoorie Biobank^2^ and the Taiwan Biobank^3^ have contributed significantly to epidemiological and public health studies in these regions. Health patterns in India (which makes up 18% of the world population) significantly influence global disease burden estimates, yet the country remains under-represented in international health research.

India’s extraordinary diversity presents unique challenges for health research. With more than 4,600 population groups and extensive endogamy over millennia^4^, genetic and phenotypic variation can align with population scales rather than administrative boundaries. Existing large-scale studies (Table S1) – including the National Family Health Survey (NFHS)^5^, Health Management Information System (HMIS)^6^, District Level Household Survey (DLHS)^7^, the National Noncommunicable Disease Monitoring Survey (NNMS)^8^, and the PhenomeIndia Study^9^—primarily focus on state-level estimates, potentially missing critical population-specific health patterns. Moreover, existing population-specific studies^10–14^ are limited in sample size, or restricted to a small number of communities, thus missing out on capturing nation-wide patterns. The lack of population-specific data means that global health models for nearly one-fifth of humanity are based on skewed or aggregated estimates. In a diverse population like India’s, patterns in small populations get masked, making it challenging to account for the genetic and environmental factors that contribute to population-specific risk .

The GenomeIndia project (GI) is a large-scale genome sequencing project from India, sequencing 10,000 healthy individuals across the country, and sampling about double this number^15^. Unlike other large surveys that sample at the state or district boundaries, the GI consortium systematically sampled 83 distinct ethnolinguistic populations across 25 states of India, accounting for both genetic diversity and population census size. Our study provides the first major assessment of phenotypic diversity across India’s ethnolinguistic landscape. Unlike samples present in electronic health records, GI samples have been collected from populations in community settings, thus reducing ascertainment bias^16^. While not large enough to direct prescriptive measures, this database is a valuable observational dataset to ascertain the health status of Indians, identifying regions and populations at higher risk for various disorders.

## Research in Context

### Evidence before study

India’s public health statistics have been characterized primarily through large national surveys that operate at the state or district level, or small ethnicity-specific surveys. These have documented a large variation in estimates of disease prevalence and burden underscoring the need for larger representation and better sampling design. Compared to global estimates, studies revealed higher burden and earlier age-of-onset for various diseases, particularly metabolic phenotypes.

Awareness and treatment gaps have been documented for hypertension and diabetes, but remain poorly characterised for dyslipidemia. Existing studies of tribal populations are small, geographically restricted and capture limited biochemical phenotypes. No large-scale study has systematically characterised metabolic phenotypes across India’s ethnolinguistic landscape

### Added Value

This is the first study to catalogue phenotypic variation across India’s ethnolinguistic diversity at scale, analysing 17,777 individuals from 81 anthropologically defined population groups spanning 25 states and union territories, sampled through a systematic community-based design. We document near-universal biochemical abnormality (95% of participants) driven primarily by an atherogenic lipid profile, and quantify awareness gaps that are more severe than previously recognised. This catalogue of population-level variation provides a reference for population-informed epidemiology studies in India. To make these findings accessible beyond specialist researchers, we have created a publicly-available interactive dashboard (https://ibdc.dbt.gov.in/gipheno/) for exploration of population-specific health patterns, trait distributions, and inter-trait correlations in GI.

### Implications of all the Available Evidence

This preliminary evidence lays the groundwork for more nuanced representative, longitudinal studies of population-specific health risk in India. We document a substantial burden of metabolic disease and large awareness gaps in the country, with the need for specific population-informed strategies for public health. These findings have direct relevance for South and Southeast Asia broadly, where similarly diverse populations within national boundaries may have fundamentally different risk profiles masked by aggregate estimates, and where some communities may have high burdens with limited diagnostic reach.

## Results

### Cohort characteristics of GenomeIndia

GI represents a unique dataset of socio-demographic, anthropometric, and blood biochemistry data (‘phenotype’ data) from 17,777 subjects across 81 ethnolinguistic population groups (from the 83 groups in GI that have phenotype data available) spanning 25 states and union territories, including several isolated populations which have been understudied (Figure 1a). To appropriately capture the genetic diversity of the complex Indian population, samples were collected across the four major language families (Indo-European, Dravidian, Austro-Asiatic, and Tibeto-Burman) across major geographic regions (which we grouped as 13 biogeographic regions as described in Subramanian et al 2026^17^) while accounting for the population census size; details of sample selection are described in Bhattacharyya et al (2025)^15^. In our analyses, consistent with previous GI work, we looked for patterns that differ between groups defined by linguistic group and tribal status: Indo-European tribes (IE_Tribe), Indo-European non-tribes (IE_NonTribe), Dravidian tribes (DR_Tribe), Dravidian non-tribes (DR_NonTribe), Austro-Asiatic tribes (AA_Tribe), Tibeto-Burman tribes (TB_Tribe), Tibeto-Burman non-tribes (TB_NonTribe), and Continentally Admixed Outgroup (CAO), a single population with known African ancestry.

**Figure 1.**
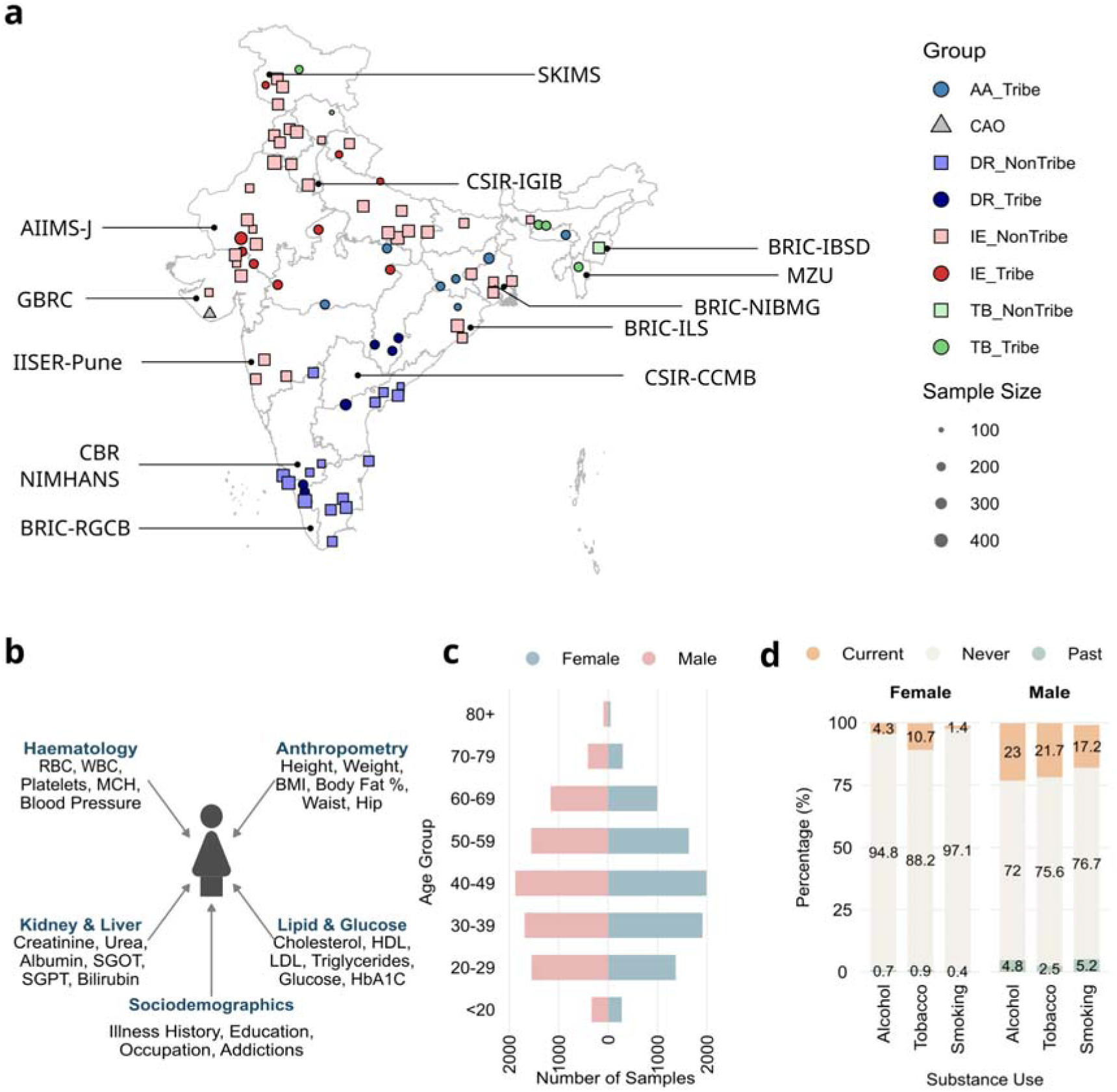
Summary statistics of the GenomeIndia (GI) phenotype dataset. A: Location and sample size of the 81 GI populations with phenotype data. Size of the coloured shapes indicates sample size. The sample collection centres and their approximate locations are indicated with labels. B: Overview of phenotype data collected for the GI cohort. C: Age pyramid for men and women (N = 17,184 individuals with available age data) D: Distribution of the prevalence of drinking alcohol, chewing tobacco and smoking for men and women.

For each individual, 67 variables were recorded (Figure 1b), of which 48 have < 25% missingness. These include 21 blood biochemistry, 19 sociodemographic, and 8 anthropometry variables (Table S2). Sample sizes with available phenotype data vary from 85 to 410 per population, with a median of 219 samples per population. 7,648 samples (43·02%) have blood biochemistry variables measured at the fasting state. Each population is encoded by an identifier that includes the linguistic group, the biogeographic region, and the tribal status of the population, as described before^17^. The dataset also contains 244 trios that enable future studies of within-pedigree correlations (Supplementary Material).

Women make up 49·38% of this dataset; the fraction of women in each population ranges from 22·01% to 75·89%. Ages span 18-97 years with a median of 43 (Figure 1c), with population-specific medians ranging from 21-57·5 years (Figure S1). 27·45% of people self-report ever having an illness or surgery, and 17·19% report being on medication. The most common medications are for hypertension and diabetes (33·96% and 22·21% of individuals on medication, respectively). There are marked sex differences in lifestyle factors: men are 12-fold more likely to smoke, two-fold more likely to consume tobacco, and five-fold more likely to consume alcohol compared to women (Figure 1d). Summary statistics for 21 selected variables (13 blood biochemistry variables with the least missingness and eight anthropometric variables of interest) are in Table 1 (Table S2 lists missingness rates for every variable).

**Table 1:** Baseline characteristics of GI samples.

| Variable | Female<br>N = 8,776<sup>1</sup> | Male<br>N = 8,998<sup>1</sup> | p-value<sup>2</sup> |
| --- | --- | --- | --- |
| Age (years) | 43.27 (14.31) | 43.99 (15.69) | 0.024 |
| Rural/Urban residence |  |  | <0.001 |
| Rural | 4,028 (49%) | 3,680 (44%) |  |
| Urban | 4,178 (51%) | 4,692 (56%) |  |
| Body Mass Index (kg/m<sup>2</sup>) | 24.28 (4.94) | 24.10 (4.34) | 0.2 |
| Height (anthropometry) (cm) | 153.65 (7.59) | 166.71 (7.81) | <0.001 |
| Weight (anthropometry) (kg) | 57.70 (13.13) | 67.02 (13.58) | <0.001 |
| Systolic Blood Pressure (anthropometry) (mmHg) | 128.09 (20.40) | 132.04 (18.83) | <0.001 |
| Diastolic Blood Pressure (anthropometry) (mmHg) | 82.46 (11.33) | 84.28 (11.73) | <0.001 |
| Waist Circumference (anthropometry) (cm) | 85.28 (13.24) | 87.90 (11.65) | <0.001 |
| Hip Circumference (anthropometry) (cm) | 94.69 (11.97) | 93.82 (9.94) | <0.001 |
| Body Fat Percentage (anthropometry) (%) | 33.87 (7.06) | 25.69 (7.85) | <0.001 |
| HDL Cholesterol (mg/dL) | 47.58 (11.48) | 43.10 (11.42) | <0.001 |
| LDL Cholesterol (mg/dL) | 108.83 (33.95) | 108.05 (34.84) | 0.4 |
| Triglycerides (mg/dL) | 148.74 (79.28) | 169.80 (90.96) | <0.001 |
| Total Cholesterol (mg/dL) | 180.74 (42.06) | 179.88 (43.21) | 0.4 |
| Haemoglobin (g/dL) | 12.14 (1.54) | 14.27 (1.69) | <0.001 |
| Aspartate Transaminase (U/L) | 27.77 (12.66) | 32.27 (14.99) | <0.001 |
| Alanine Transaminase (U/L) | 24.17 (13.31) | 32.52 (17.40) | <0.001 |
| Creatinine (mg/dL) | 0.73 (0.17) | 0.91 (0.19) | <0.001 |
| Glycosylated Haemoglobin/HbA1c (%) | 5.69 (0.87) | 5.78 (0.97) | <0.001 |
| Urea (mg/dL) | 21.99 (8.19) | 23.16 (8.09) | <0.001 |
| Alkaline Phosphatase (U/L) | 95.32 (33.98) | 96.74 (33.06) | <0.001 |
| Total Bilirubin (mg/dL) | 0.50 (0.25) | 0.62 (0.29) | <0.001 |
| Direct Bilirubin (mg/dL) | 0.15 (0.09) | 0.19 (0.12) | <0.001 |

1 Mean (SD); n (%)
2 Wilcoxon rank sum test; Pearson's Chi-squared test

### Phenotype variation across Indian populations

All recorded phenotypes show significantly different distributions between populations, and also between linguistic groups and biogeographical regions. These differences remain after accounting for confounding variables like age, gender, and sampling center. Phenotype prediction significantly improves when adding population as a predictor to models using age, gender and state. The mean increase in adjusted R^2^ is 7·4% (Table S3). Among metabolic variables, waist circumference has the largest gain, with R² increasing by 10·7%. Populations in the state of Odisha illustrate this pattern: different populations in the same state have significant differences in several metabolic characteristics (Figure 2a, Figure S2a for individuals without medication). Previous studies that capture data at the state resolution, like NFHS, miss this pattern of ethnicity-structured variation.

**Figure 2.**
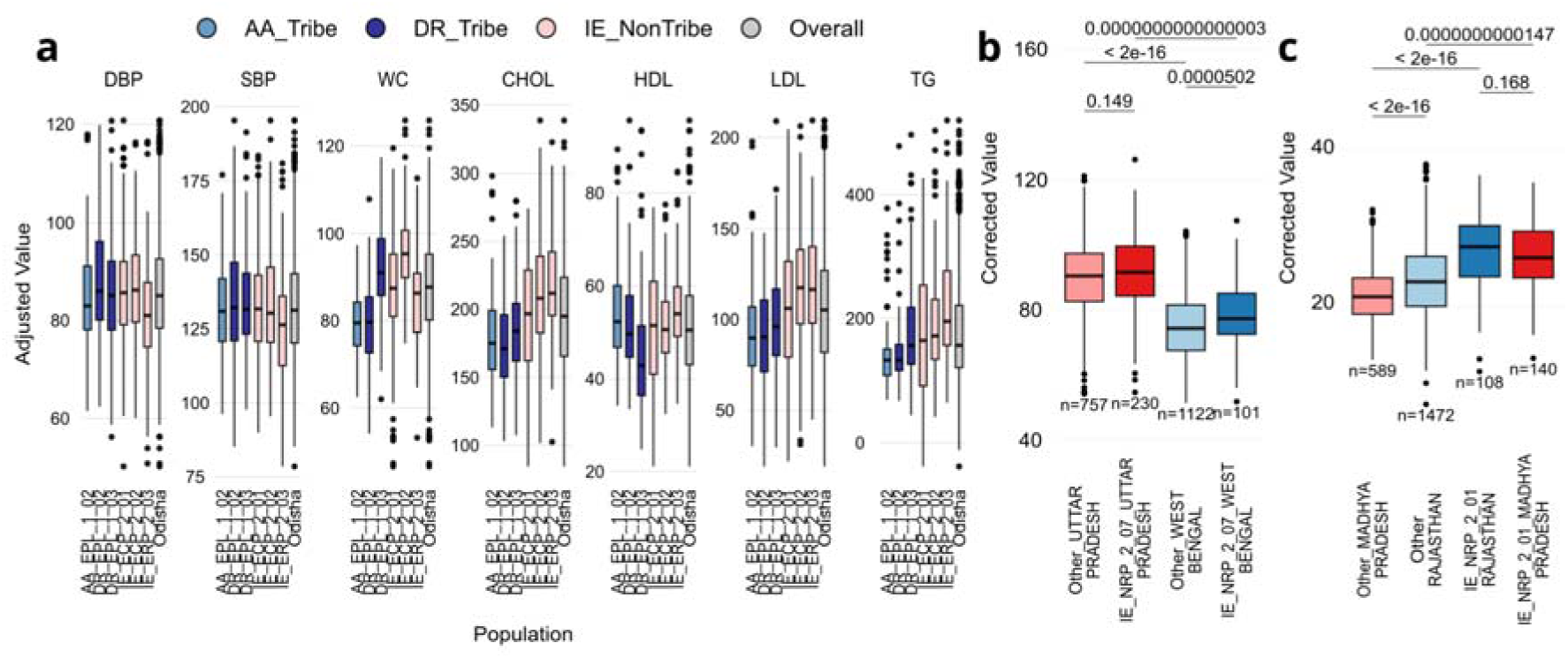
Phenotype variation across Indian populations a: Variation in selected indicators for the six populations sampled from Odisha, corrected for age, gender, sampling center and fasting status, compared against the overall state distribution (DBP - diastolic blood pressure, SBP - systolic blood pressure, WC - waist circumference, CHOL - total cholesterol, HDL - HDL cholesterol, LDL - LDL cholesterol, TG - triglycerides) b: Illustrative example of the same population IE_NRP_2_07 (in deep colors) spread across two states (each state is in a different color), showing a stronger geography effect in waist circumference c: Illustrative example of the same population IR_NRP_2_01 (in deep colors) spread across two states, showing a stronger population effect in BMI

Eighteen ethnolinguistic groups were sampled across two states (n ≥ 30 per state), allowing us to ask whether phenotypic distributions within the same population differ by geographic location. The degree of geographic variation is highly trait-dependent (Figure S2b, Table S4). Liver enzymes, urea, and direct bilirubin show significant and strong state differences in multiple populations, which may reflect a sensitivity to environmental exposures. In contrast, BMI distributions are largely similar across sampling locations for most populations. Notably, waist circumference shows significant geographic differences in several populations despite its close relationship with BMI.

### Systemic burden of metabolic risk across Indian populations

Strikingly, 95·02% of the population (16,892 of 17,777 samples) has abnormal values for at least one of 14 health parameters (those below 20% missingness) (Table S5) (Figure 3a; Table S2 for reference ranges and thresholds). This burden remains consistent after excluding medicated individuals and non-fasting samples (Figure S3a, S3b, Table S6).

**Figure 3.**
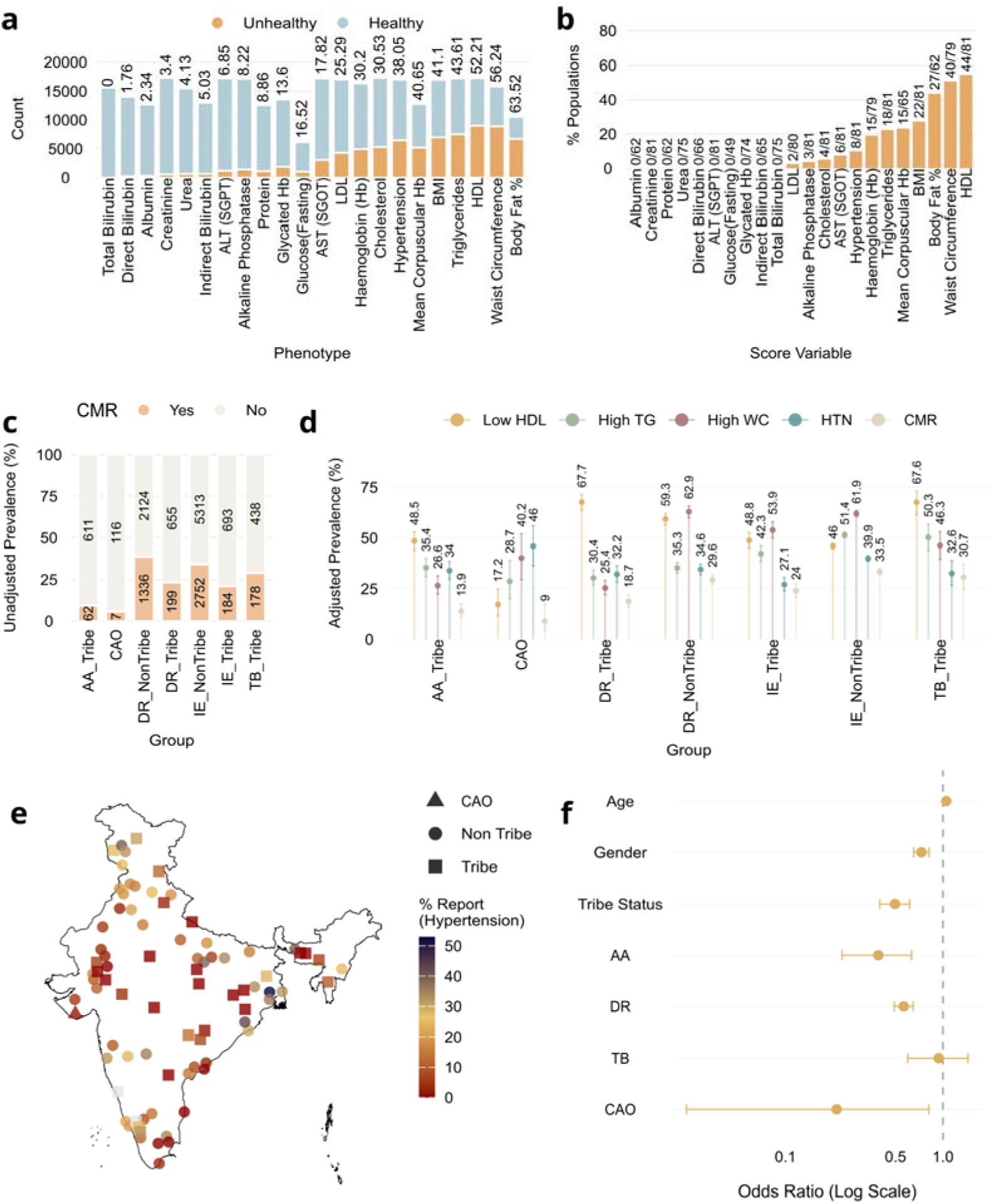
Prevalence of phenotypic abnormalities in the sampled populations. a: Counts of healthy and unhealthy samples for 22 phenotypes sorted by percentage of unhealthy values (blood biochemistry and anthropometric, excluding blood cell counts) b: Number of populations that have 50% or more samples flagged as unhealthy for the 22 phenotypes in a. c: Counts of individuals with and without clustered metabolic risk in each group d: Age and gender-adjusted risk for each risk factor in the clustered metabolic risk symptoms e: Geographical distribution of proportion of hypertensive individuals who report it f: Odds ratio of awareness (reporting/medication) among hypertensives, adjusting for age, gender, ancestry group and tribal status (reference set as IE, Non-Tribe, Female)

The high prevalence is driven by an atherogenic lipid profile^18^: 52·21% (8,993 of 17,226 samples) have low high-density lipoprotein cholesterol (HDL), and 43·61% (7,498 of 17,194) have high triglycerides. 44 populations have more than 50% of individuals with low HDL (Figure 3b, figure S3c for non medicated, S3d for fasting). When looking at fasting samples specifically, the low HDL prevalence does not change, and the high triglycerides prevalence is 38%, confirming that the lipid burden is not an artifact of fasting status. As 57% of samples were collected in the non-fasting state, primary analyses include both fasting and non-fasting samples; studies have shown that non-fasting lipid levels can capture cardiovascular risk as well as fasting values^19^.

To illustrate the difference in burden between populations, we focused on four of the five risk factors that define metabolic syndrome with low missingness in our dataset (low HDL, high triglycerides, high waist circumference, and hypertension), and classified individuals with at least three risk factors as having ‘clustered metabolic risk (CMR)’^20^. Individuals in the ‘CAO’ group have the lowest adjusted prevalence (5·7% raw, 9% adjusted for age, gender and sampling center) of clustered metabolic risk (Figure 3c), and the Indo-European non-tribe and Tibeto-Burman tribal groups have the highest adjusted prevalence (34·1% and 28·9% raw respectively, and 33·5% and 30·7% respectively, adjusted). These patterns remain when including only those individuals who are not on medication. (Table S7A, S7B, Figure S4a). Metabolic syndrome quantified in those samples with measured fasting glucose values follow the same patterns as ‘clustered metabolic risk’ (Table S8A, Table S8B).

These four risk factors vary significantly across the seven groups. In non-tribal populations from IE and DR language groups, high waist circumference is the most prominent risk factor, while in the tribal populations from IE, DR, AA and TB groups low HDL dominates. In CAO, hypertension appears most crucial (Figure 3d, Figure S4b).

GI populations also vary significantly in the prevalence of abdominal obesity, which has been extensively studied in the Indian context for its distinct distribution^21,22^. Prevalences range from 25-27% in Dravidian tribes and Austro-Asiatic tribes respectively to 63% in Dravidian non-tribal populations (Figure 3d, Figure S4c). BMI, a related trait, varies similarly; while Austro-Asiatic tribes primarily have a large fraction of underweight individuals, non-tribe populations show a high fraction of overweight individuals (Figure S4d). 22 out of the 81 populations show a double burden in terms of having 10% or more underweight and 20% or more overweight individuals within the same population.

Diabetes prevalence in GI (18·41%) is comparable with INDIAB^22^ (21·1%), but is four-fold higher than the NFHS-5 estimates (4·39%; Table S4); the difference with NFHS reduces when prevalence is estimated from an age-matched subset of individuals < 50 years old. Hypertension prevalence in GI (41·51%) is higher than in INDIAB (35·5%) and NFHS (19·84%), even after matching for age (Table S9).

These data suggest that abnormality is not an outlier condition, but a baseline characteristic of most population groups in India. This raises questions about the validity of Western-derived reference intervals in Indian populations, which can lead to systematic over-diagnosis, or whether India is facing a large metabolic burden^23^.

### Burden of undiagnosed disease

A large fraction of people with abnormal metabolic values do not self-report any illness or being on any medication, reflecting and reinforcing the burden of untreated and undiagnosed complex disorders^13,24–26^. For instance, out of 6,476 individuals whose recorded blood pressures are in the hypertensive range, only 1,141 (17·6%) either self-report hypertension or are on antihypertensive, heart-related, anticoagulant or unknown medication (Figure 3e). Individuals with diabetes (717 reporting out of 2,677, 26·8%) are the most likely to be diagnosed, followed by hypertension (17·6%) and then dyslipidemia (Figure S5a,b,c), which has a reporting fraction of 2·2% (214 reporting out of 9,853). The CAO population has the lowest reporting fraction for all three, pointing to potential challenges in healthcare access in this community (Figure S5d). Tribal groups show lower awareness compared to non-tribal groups (Figure 3f). Differences in reporting fractions by populations could reflect disparities in access to public health, and point to areas most in need of increased awareness and medical resources.

### Life-course patterns: interactions with age and gender

Age- and sex-related trajectories in GI broadly recapitulate patterns documented in European cohorts, but with some population-specific patterns. The top few principal components of blood biochemistry data within each population group partially or completely separate male and female participants even after adjusting for age (Figure S6), suggesting sexual dimorphism. Indeed, women have higher body fat^27^, lower haemoglobin levels^28^, lower serum creatinine^29^, and lower HbA1c^30^ and higher HDL levels^31^ (Table 1). Haemoglobin shows the most consistently sexually dimorphic trajectory of any measured trait, with significant sex differences at every age bin. While women’s haemoglobin levels remain relatively stable with age, men show a steady decline across the life course (about 0·22 g/dL decreased haemoglobin per decade increase in age), consistent with prior reports^32,33^.

The relationship between metabolic phenotypes and age also differs between men and women. Given the effect of medication on health parameters, we restrict analyses to individuals not on any medication (Figure 4a, Figure S7a for all individuals). All lipid markers show significantly different trends with age for men and women (defined by the interaction term between gender and age group), as shown before^31,34,35^ (p < 0·01; Table S10A, Table S10B). Mean total cholesterol increases with age up to ∼50 years, after which it declines gradually in men, while women show an elevation at ∼45 years old, which remains elevated. Previous studies have attributed this trend in women to menopause-related changes^36^; the reasons underlying the decline for men is not clear. For triglycerides, younger men show a higher mean level compared to women until age 50, after which the trend reverses. In women, triglycerides increase until a decade older–age group 60-69, a reversal point earlier than most existing studies^31,37^ apart from the Dutch Lifelines study (reversal around 40-44 years of age^34^).

**Figure 4.**
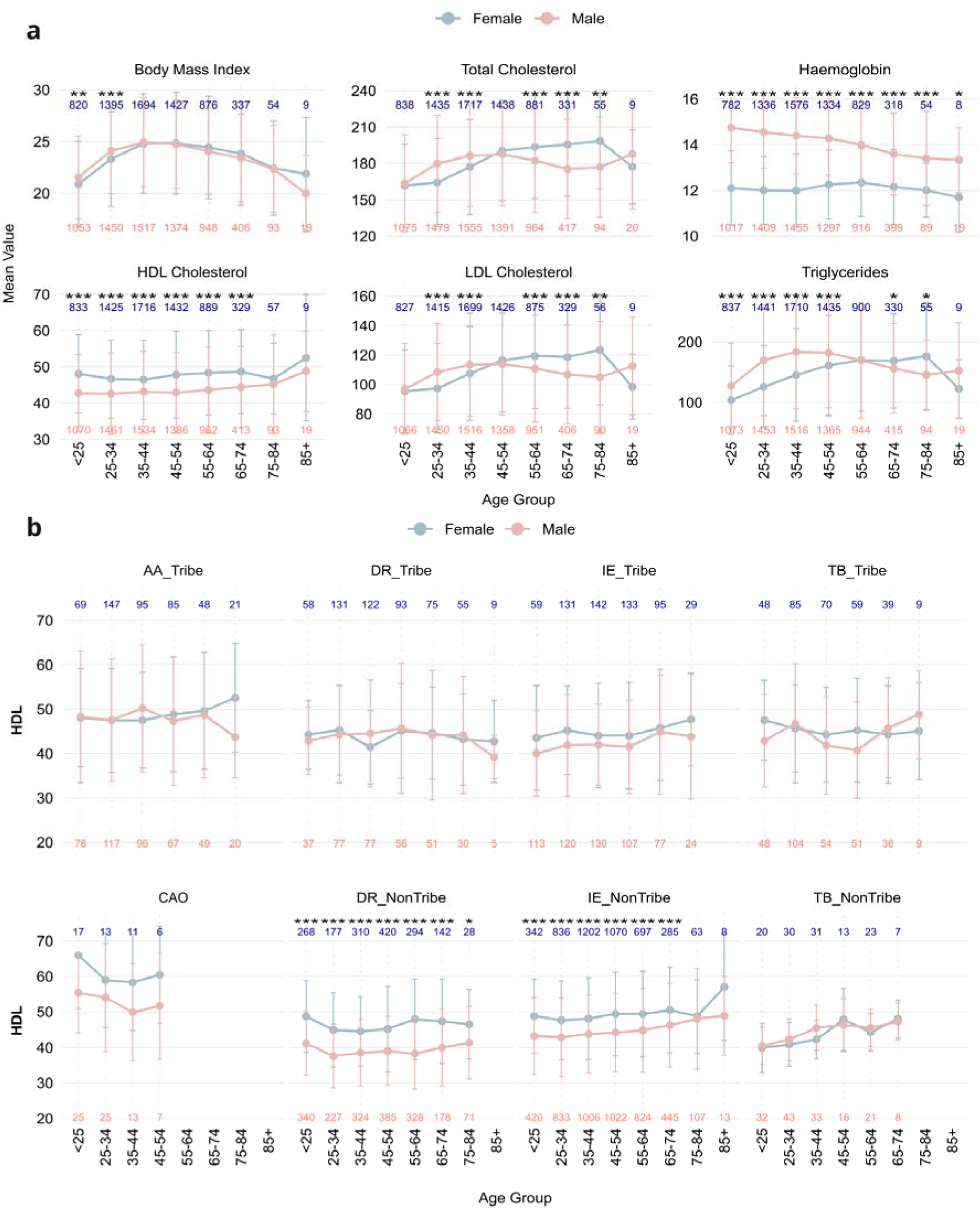
Life course patterns of biomarkers among male and female participants. A: life-course trends for a few selected biomarkers between male and female participants. B: life-course trends for HDL cholesterol for the eight groups, displayed for bins with 5 or more male and female samples each (BH-adjusted p<0·05 is significant. Adjusted p < 0·001 - ***, adjusted p < 0·01 - **, adjusted p < 0·05 - *)

HDL and triglyceride levels significantly differ between men and women, unlike LDL and total cholesterol levels (Table S10A, S10B). These trends remain robust across alternative choices of age binning (Figure S7b). Notably, while higher female HDL levels are consistent across non-tribal ancestry groups (a pattern also reported previously in European and Indian populations^38,39^), this pattern is absent in tribal populations across all language families (AA, DR, IE, and TB), suggesting that the factors driving sex differences in HDL metabolism are different in tribal groups (Figure 4b, Figure S8a, S8b).

## Discussion

We present the first large-scale phenotypic characterisation of India’s ethnolinguistic diversity, documenting systematic health patterns across 81 population groups. Our sampling design reveals health differences which national averages miss. Our findings demonstrate that ethnolinguistic identity is a significant predictor of metabolic health outcomes, independent of geographic location, age, and sex. This suggests that a population’s genetic ancestry and socio-cultural environment both shape health risk in ways that administrative boundaries fail to capture. Alongside this variation, we document a high and near-universal burden of metabolic abnormalities, and striking gaps between disease prevalence and clinical awareness, with major implications for public health policy in India and similarly diverse populations.

The dataset reveals striking variation in metabolic risk across ethnolinguistic groups; clustered metabolic risk prevalence ranges from 9% (adjusted for age, gender and sampling center) in CAO populations 33.5% (adjusted) in Indo-European non-tribal populations, approximately a four-fold difference. This variation is not captured by state-level surveys which aggregate across populations with fundamentally different risk profiles, specifically failing for the tribal populations. Our findings suggest that ethnolinguistic identity be considered in population-specific risk assessment frameworks.

A striking pattern emerging from life-course analysis is that the pattern of higher HDL levels typically observed in women compared to men are absent in tribal populations across all language families. In non-tribal groups, women consistently show higher HDL than men across all age bins, mirroring patterns in European and urban Indian cohorts. This sex difference in HDL disappears in tribal populations, suggesting that the factors driving it may be systematically different in these communities. South Asian women in general already have lower absolute HDL levels than European women^38–40^; the further loss of this sex difference in tribal groups compounds an already high cardiovascular risk in these populations. Whether this reflects a genuine biological difference, a lifestyle effect, or differential exposure to environmental factors warrants targeted investigation.

95% of participants had abnormal values for at least one biochemical or anthropometric parameter. The near-universal prevalence likely reflects two factors: a genuinely high metabolic burden in Indian populations driven by an atherogenic lipid profile, and systematic misclassification arising from reference intervals derived from European populations applied to South Asians. These explanations are not mutually exclusive. The 95% figure could therefore be interpreted as an upper bound on true disease burden. However, the burden is unlikely to be merely an artefact of incorrect thresholds; elevated waist circumference (which was assessed using a South-Asian specific cutoff) has a high prevalence across many groups. To quantify the true extent of metabolic burden, establishing population-specific reference intervals for Indian populations must be an urgent priority.

The awareness gap we document represents an actionable public health issue. Of 6,476 individuals with blood pressure in the hypertensive range, only 1,141 were aware of or being treated for their condition. For dyslipidemia, awareness was near-absent at 2·2%. These gaps were most pronounced in tribal populations, where barriers to healthcare access are well-documented^41,42^. This is not simply a problem of disease burden, but also a problem of detection. This has direct implications for public health priorities in India and across Southeast Asia, where similarly underserved populations may face comparable barriers to diagnosis and treatment. Targeted screening programmes, particularly for hypertension and dyslipidemia in tribal and rural communities, are an immediate and tractable intervention.

Though this is one of the first datasets from the country that captures such a broad ethnolinguistic range, it still remains a first step. The cross-sectional design precludes assessment of whether observed biomarker patterns predict clinical outcomes; this would require longitudinal data. Moreover, smaller communities require more extensive sampling before confident estimates of prevalence can be established. Self-reported data for variables such as medication status, history of illness and smoking/tobacco/alcohol usage introduces an additional degree of uncertainty to any downstream interpretation. Finally, there could be a participation bias, for example, of health-conscious volunteers, or people reluctant to reveal their disease status. Despite these limitations, the consistency of findings across robustness checks and the breadth of population coverage strengthen confidence in the overall patterns reported.

This study establishes the first comprehensive phenotypic map of India’s ethnolinguistic diversity, revealing a landscape of health risk that is highly structured across populations. The findings demonstrate that state-level health data obscures variation that is epidemiologically meaningful. Addressing India’s metabolic health crisis requires moving beyond administrative boundaries toward population-specific surveillance, reference intervals, and targeted interventions. The GI phenotypic dataset, freely accessible through an interactive dashboard (https://ibdc.dbt.gov.in/gipheno/), provides a foundational resource for this transition.

## Methods

### Sample collection strategy

Efforts to educate and sensitize GI representatives and to ensure standard collection protocols are described in the supplementary material. Detailed socio-demographic data, anthropometric and blood biochemistry data were collected as the phenotype data. To ensure uniformity in data collection, detailed SOPs were put into place for each category of phenotype data. For blood biochemistry data, registered diagnostic centres processed the samples to maintain quality and adherence to regulatory standards. In some cases, hospital-based centres processed samples in-house. All field data (sociodemographic and anthropometric) were entered using the Open Data Kit (ODK), which was directly linked to the server at the Centre for Brain Research (CBR), the coordinating centre, to ensure centralized data collection and management.

Ethics and information sheets: All participating centres obtained ethical approval from their respective Institutional Ethics Committees. The 13 sample collection centres obtained informed written consent from all participants before recruitment into the study, in accordance with the ethical guidelines outlined by the Indian Council of Medical Research (ICMR). Consent forms were provided in both regional languages and English to ensure that participants clearly understood the purpose, procedures, risks, and benefits of the study in a language they were comfortable with, before agreeing to participate.

### Data integration and preprocessing

All initial data cleaning steps are described in the supplementary methods. After data cleaning, we removed (marked NA) values that are out of range, or outside observed ranges; we report the thresholds used to filter out values for each variable in Table S2. We limited filtering to very conservative thresholds to allow users of this dataset to set their own thresholds for each variable. The final dataset includes 67 variables, which comprise of all 40 anthropometric and socio-demographic variables, 25 blood biochemistry variables with missingness < 30%, and random and fasting glucose (2 variables, which had higher missingness of up to 65%, but are of interest to many researchers) (Table S2). We have used self-reported gender for all the participants with available phenotype data.

For all statistical analyses in this study, we performed one more round of statistical tail-aware outlier removal. For each variable, we measured the skewness, and marked as NA those values that are more than 3SD above or below the mean for the tail that is more skewed. To demonstrate the robustness of this outlier removal method, we repeated all analyses with outlier removal using the extreme 0·5 percentile of values. All patterns discussed in the results remained consistent. To maintain reproducibility, we performed all curation and data pre-processing using R scripts (R version 4.3.3).

### Clinical definitions

Rules used to define hypertension, dyslipidemia, diabetes, metabolic syndrome, and clustered metabolic risk use common guidelines, and are described in the supplementary methods.

## Statistical analysis

1. Baseline characteristics of the study individuals: We assessed the difference in summary statistics between men and women. For continuous variables, we reported the mean and standard deviation. For rural/urban residence, we reported the number and percentage of samples in each group. For continuous variables, the statistical difference between men and women is reported using p-values from a Wilcoxon rank-sum test; for categorical variables, we used the Pearson’s Chi-square test.
2. Assessing increase in the performance of phenotype prediction on adding population: For each of the 37 quantitative variables in our dataset, we fitted two OLS linear regression models, Model 1: Phenotype ∼ age + gender + state and Model 2: Phenotype ∼ age + gender + state + population. Gender, state and population were encoded as categorical factors. The increase in the prediction accuracy was quantified by the difference in adjusted R^2^ between model 2 and model 1. We reported both the unadjusted and adjusted R^2^ values. The p-values were calculated with an F-test between the two models. We used a Benjamini-Hochberg adjusted p-value threshold of 0·05 to report significance.
3. Phenotypic variability within multiple populations present in a state: We used a linear regression model to correct for variation in the four lipids and the remaining three phenotypes responsible for CMR, adjusting for age, gender, sampling center and fasting status. We added the residuals to the population mean to get the corrected values on the same scale as the original, and compared the distributions for each population in the state.
4. Assessing phenotypic association with population identity or states: To determine the extent to which population or state explain the variation in phenotype, we ran two regression models Phenotype ∼ Age + Gender + State/Population. The difference in R-squared of these two models with the base model Phenotype ∼ Age + Gender gave the extent of variation explained by each factor.
5. Calculation of risk for clustered metabolic risk factors: We used a generalised logistic regression model, with the presence or absence of clustered metabolic risk factors (low HDL, high triglycerides, high waist circumference, hypertension) as the response variable, and ethnolinguistic group (structured as ancestry_tribe/nontribe), age, gender, and sampling center as the predictors to get adjusted prevalences/risks for each of the risk factors, and CMR as a whole. We repeated the same analyses for the subset of individuals not on medication.
6. Comparing prevalence of diabetes and hypertension with other pan-India datasets: To compare the prevalence of diabetes and hypertension with other large pan-India studies (NFHS-5, ICMR-INDIAB and LASI), we calculated the prevalence of diabetes and hypertension in the GI dataset according to the criteria described by these studies. We also calculated prevalence for all GI individuals under 50 and a gender ratio-matched subset to keep statistics comparable to NFHS-5. For comparing statistics to LASI, we took the subset of GI individuals aged 45 or more.
7. Underreporting of conditions among groups: To assess the odds ratio of underreporting among various groups, we ran a logistic regression model. The dependent variable was awareness (1/0), defined by reporting a condition, which was regressed against Age + Gender + Ancestry Group + Tribal status for all the individuals who either biomedically had or reported diabetes, dyslipidemia and hypertension. For all the three conditions, IE, Non-Tribe Female was taken as the reference group and odds ratios calculated compared to this reference.
8. Life-course patterns: We binned samples into eight age groups (<25, 25-34, 35-44, 45-54, 55-64, 65-74, 75-84 and >85). To assess differences in the trend with age, gender and the interaction of age and gender, we performed ANOVA in the format ANOVA(Phenotype ∼ Age Group * Gender). For each biomarker, we determined significant differences in the means between males and females using a t-test, correcting for multiple testing with the Benjamini-Hochberg correction (significance defined as adjusted p-value < 0·05.)

## Data Availability

De-identified individual-level data are available through controlled access from the Indian Biological Data Center (IBDC) to protect participant privacy (https://ibdc.dbt.gov.in/data_access/). Population-level data visualizations are freely available through the interactive dashboard at https://ibdc.dbt.gov.in/gipheno.

## Acknowledgements

We thank all participants who consented to providing their samples for this project. We acknowledge funding by the Department of Biotechnology (DBT), Ministry of Science and Technology, Government of India (BT/GenomeIndia/2018). We acknowledge the current and past leadership of all participating institutes in the GI Project. The authors thank the support provided by the computational facilities of the institutes. We acknowledge the use of generative AI and large language models (Claude Sonnet 4.6) in refining the language of the manuscript; however, the responsibility of the final content lies with the authors.

## Author Contributions

Debasrija Mondal: Data Curation, Formal Analysis, Writing–original draft, editing and reviewing, Dashboard - conceptualisation and building. Chandrika Bhattacharyya: Investigation, Data Collection and Curation, Formal Analysis, Writing – review & editing.

Dolat S Shekhawat, Nayan G Tada, Tanuja Rajial, Arun Sree Parameswaran, Deepak Jena, Sudeshna Datta, Mamuni Swain, Sudarshan Jena, Adyasha Mishra, Soumendu Mahapatra, Shijulal Nelson Sathi, Mahabub Alam, Azad Ali, Parveena Choudhury, Poulomi Ghosh,Shobha Anilkumar, Divakar Ashwath, Mohana Chithimmaiah, Shafeeq K Shahul Hameed, Rajesh Gunasegaran, Neha Singh, G Mala, Tiyasha De, Shahrumi Reza, Ankit Mukherje^e^, Bhumika Prajapati, Bhagirath Dave, Silvia Yumnam, Kshetrimayum Vimi, Gurumayum Nikesh Sharma, Ajay Malik, Ranjan Jyoti Sarma, Andrew Vanlallawma, Doddaladka Krishnayya Samartha, Tejaswini S G, Paranthaman V Kavya: Investigation, Data Collection and Curation.

Devashish Tripathi: Formal Analysis

Sanjay Deshpande: Dashboard - conceptualisation and building

Kuldeep Singh, Nanaocha Sharma, Sunil K Raghav, Chaitanya Joshi, Madhvi Joshi, Santosh Dixit, Mayurika Lahiri, H. Lalhruaitluanga, Lal Nundanga, Nachimuthu Senthil Kumar, Ganesan Venkatasubramanian, Naren P Rao, Venkataram Shivakumar, E V Soniya, Mohd Ashraf Ganie, Imtiyaz Ahmad Wani, Bratati Kahali, Divya Tej Sowpati, Mohammad Faruq, Sridhar Sivasubbu, Vinod Scaria, Arindam Maitra, Kumarasamy Thangaraj, Vijayalakshmi Ravindranath: Investigation, Data Collection and Curation, Conceptualization.

Ganganath Jha, Prathima Aravind, Karthik Tallapaka, Abdul Jaleel, Suman Paine: Conceptualization, Investigation, Data Collection and Curation, Formal Analysis, Writing – review & editing (equal contribution).

Analabha Basu, Shweta Ramdas: Conceptualization, Investigation, Data Curation, Formal Analysis, Writing – original draft, Writing – review & editing, Supervision (equal contribution, co-corresponding authors).

