## Supplementary Material for "Capturing India’s phenotypic diversity: Health insights from the GenomeIndia project"

### **Table of Contents**

**Table of Contents	1**

**Section 1: Supplementary Methods	2**

**Section 2: Trios in the GI cohort	5**

**Section 3: Search Strategy for existing literature	6**

**Section 5: Supplementary figures	8**

Figure S1	8

Figure S2	9

Figure S3	10

Figure S4	11

Figure S5	12

Figure S6	13

Figure S7	14

Figure S8	15

**References	16**

### **Section 1: Supplementary Methods**

*Sample collection strategy*

All representatives from GI sample collection centres participated in a workshop titled “Population Identification, Field work and Sample Collection for GenomeIndia” held at the Institute of Genomics and Integrative Biology in March, 2020. This workshop was led by expert anthropologists and professors with prior experience in population genomics studies in India. Key focus areas included population identification, and approaching people from different Indian population groups for participation in the GI project. Hands-on practice sessions were also conducted.

Tribal groups were mapped to specific, dedicated regions, while isolated and larger populations were identified across both rural and urban settings. Tribal populations were approached only after obtaining the necessary permissions from the respective state governments. Isolated populations and larger populations were approached through local leaders, community representatives, and through relevant organizations. After 83 unique population groups were identified to maximize diversity, they were allocated to 13 partnering centres for sample collection based on their geographical locations. Field visit teams from these centres visited the villages, organizations, and specific locations to enrol participants from each population group. For isolated regions, carefully planned collaborations with local healthcare institutions helped in streamlining data collection and improving accessibility.

Before the collection of data, outreach and community engagement programmes were conducted to emphasize the importance of genetic research, clarify the study’s objectives, and assure participants about data confidentiality. Common Standard Operating Procedures (SOPs) were developed and consistently followed for the collection of biological samples and phenotypic data, including demographic and anthropometric information.

*Data cleaning and preprocessing*

We reformatted all the blood biochemistry files to standard formats, and standardized column names for all variables to a common set of names. We rescaled all variables across centres to a standard set of units. Following this, we collated data from all centres, and merged this with the anthropometric data for each sample.

We also cleaned values in columns containing factor variables into a set of uniform labels.These included correcting typos in state names, recoding the smoking, chewing tobacco and drinking alcohol status to a current/past/never coding system, encoding the monthly family income to a set of seven scores (1, 2, 3, 4, 6, 10 and 12), and encoding marital status to never married, currently married, divorced or separated, and widowed.

Three columns–date of birth, approximate age, and age on interview–contained age information. Values in these columns were highly correlated; 99.61% between approximate age and age on interview, 99.34% between approximate age and calculated age, and 99.75% between age on interview and calculated age. We used ‘date of birth’ (the column with the least missingness) as the default column for calculating sample age (difference between date of birth and date of interview). For samples with missing values in calculated sample age, we used values from approximate age or age on interview, as available. The final missingness for the ‘age’ variable was 3.3%.

Family income was reformatted as tiered scores to ensure uniformity across the dataset. The scores and income ranges (in INR per month) correspond to 1: ≤2,640, 2: 2,641-7,886, 3: 7,887-13,160, 4: 13,161-19,758, 6: 19,759-26,354, 10: 26,355-52,733 and 12: ≥52,734.

*Rural-urban classification*

Using the approximate location for each study subject as part of the sociodemographic data, we tallied each entry against the list of villages and towns according to the 2011 Census of India ^43^, as well as the OneFiveNine^44^ directory. Based on matching cleaned ‘village’ and ‘state’ names, we assigned samples a rural or urban classification. 1,198 samples (6.74%) remained unclassified in the end because of ambiguous location or missing precise location.

*Definition of medication type*

We classified the names of medication given according to the categories of their function, using the keywords “telmi” “telma” “telva” “bp” “olme” “olmi” “amlo” “card” “losar” “htn” “heart” “ecosprin” “pressure” and their spelling variants for available medicines for hypertension. Similarly, “glime” “metformin” “glyco” “insulin” “gluca” “sugar” “diabetes” “diabetic” were used for diabetes, and similarly other keywords were used, such that each unique medicine name gets mapped to a classification that groups them together according to function. For the medications with a name that could not be reliably classified, we marked them as ‘unclear’ in a separate variable.

*Clinical definitions*

*Hypertension:* We classified any sample with >= 140 mm Hg systolic blood pressure, or >= 90 mm Hg diastolic blood pressure, or both, as hypertensive according to the Eighth Joint National Committee (JNC 8)^3^. If either of systolic or diastolic blood pressure was NA, we marked hypertension status as NA.

*Dyslipidemia:* According to the NCEP-ATP III^4^ guidelines for dyslipidemia, we defined dyslipidemia by the presence of one or more of: high total cholesterol ≥ 200 mg/dL (hypercholesterolemia), low HDL <40 mg/dL for men, and <50 mg/dL for women (hypoalphalipoproteinemia), high LDL ≥ 130 mg/dL (hyperlipidemia), and high triglycerides ≥ 150 mg/dL (hypertriglyceridemia).

*Diabetes:* We defined diabetes by the presence of one or more of the following, according to the criteria specified by the American Diabetes Association (2010)^5^ : glycated haemoglobin ≥ 6.5%, fasting blood glucose ≥ 126 mg/dL, and random blood glucose ≥ 200 mg/dL.

*Metabolic syndrome:* We defined metabolic syndrome according to the presence of three or more of the following NCEP-ATP III^4^ criteria: 1) waist circumference ≥90 cm for men or ≥80 cm for women, 2) Triglycerides ≥150 mg/dL, 3) HDL-C < 40 mg/dL in men or <50 mg/dL in women or on specific treatment for this lipid abnormality, 4) systolic blood pressure (SBP) ≥130 mm Hg or diastolic blood pressure (DBP) ≥85 mmHg or both, or under treatment for hypertension, 5) fasting plasma glucose ≥100 mg/dL (HbA1C: >6%), or previously diagnosed type 2 diabetes mellitus.

*Clustered metabolic risk:* We defined clustered metabolic risk according to the presence of three or more of the following metabolic risk factors for metabolic syndrome (as defined above): 1) HDL-C < 40 mg/dL in men, <50 mg/dL in women, 2) triglycerides ≥150 mg/dL, 3) waist circumference ≥90 cms for men or ≥80 cm for women, and 4) systolic blood pressure (SBP) ≥140 mm Hg or diastolic blood pressure (DBP) ≥90 mmHg or both, or under treatment for hypertension, and classified individuals with at least three risk factors as having ‘clustered metabolic risk (CMR).

*Definition of underreporting of conditions*

For any particular condition, we considered awareness as either reporting the condition as part of their history of illness, or having the relevant category of medication according to our classification. For being on medication, we considered all the individuals having the relevant category of medication. To prevent underestimating the reporting and medication fractions, we also considered all the individuals who had any ‘unclear’ medication types in reporting and medication. We calculated the reporting fraction as the number of individuals having the condition and reporting it divided by the number of individuals having the condition. For statin-treatable dyslipidemia, we considered all individuals who had at least one of high LDL, high total cholesterol and high triglycerides as dyslipidemic, since the relevant category of medication was statins, which primarily lower LDL cholesterol, total cholesterol and triglycerides.

### **Section 2: Trios in the GI cohort**

The GI sequenced dataset includes 244 trios confirmed by genetic relatedness estimates. While the trio sample size only allows for exploratory analyses, we analyzed lipid (LDL, HDL, total cholesterol and triglycerides) correlations between parents and children.

After adjusting lipid levels for age, BMI, smoking status and lipid-lowering medication (fitted separately for father, mother and child), we calculated pairwise Spearman correlations for these adjusted values, comparing father-child and mother-child pairs. After adjustment and dropping missing values, we were left with 174 complete trios for LDL, 177 for HDL, 178 for cholesterol and 170 for triglycerides. Considering that the lipid traits themselves are highly correlated, we adjusted the p-values by Benjamini-Hochberg correction.

Modest but significant parent-offspring correlations were observed for all four lipid traits, consistent with the polygenic and environment-affected nature of these traits. HDL showed the strongest familial signal; the father-child correlation was 0.41, which was stronger than the mother-child correlation at 0.30. This contrasts with the other traits, where the mother-child correlation was always stronger than the father-child correlation. Moderate mother-father correlations exist for all lipid traits except triglycerides, suggesting some degree of assortative mating. (Table S11)

### **Section 3: Search Strategy for existing literature**

Tool used: Perplexity AI

Each individual result was verified by subsequent manual inspection to prevent hallucination of citations.

{

"task": "Generate table of epidemiology/public health studies in India (≥200 participants)",

"instructions": "Include national-level, regional, district, state, cohort, ethnicity, tribe level studies . Use peer-reviewed papers with DOI.",

"required_fields": [

"DOI",

"Study",

"Journal",

"Sample Size",

"Key Points"

],

"criteria": {

"temporal": "2000-2026",

"geographic_scope": "National/regional/state/district/cohort/ethnicity/tribe ",

"participant_threshold": "≥200 participants total",

"focus": "Epidemiology, Public health, Hypertension/Diabetes/Dyslipidemia prevalence, risk factors, treatment cascade, control rates",

"sources": "Peer-reviewed journals ONLY (PubMed, Lancet, BMJ, J Hum Hypertension, Indian J Med Res)",

"doi_requirement": "Must have valid DOI (no conference abstracts)",

"key_points_include": [

"Prevalence % by age/sex/urban-rural",

"Awareness/treatment/control cascade",

"Risk factors identified",

"Regional hotspots",

"Treatment gaps",

"Tribes at risk"

]

},

"examples_to_target": [

"NFHS-4 district-level analysis (BMJ Open)",

"NNMS hypertension cascade (J Hum Hypertension)",

"ICMR-INDIAB state-wise (Indian J Med Res)",

"CURES Chennai cohort (J Assoc Physicians India)",

"STEPS Tamil Nadu/Kerala",

"Ballabgarh HDSS hypertension cohorts",

"LASI Wave 1"

],

"search_hints": {

"district": ["NFHS-4 hypertension district", "NFHS-5 hypertension state"],

"cohort": ["ICMR-INDIAB", "CURES hypertension", "Kalyani cohort hypertension"],

"steps": ["STEPS Tamil Nadu hypertension", "STEPS Kerala hypertension"]

},

"output": {

"format": "Markdown table",

"sort_by": "Sample size descending",

"minimum_studies": 50,

"regional_summary": "Highlight tribes at risk + national patterns + urban-rural gradients"

},

"exclusions": [

"<200 participants",

"No DOI papers",

"Pre-2000 studies"

]

}

Date of search: March 5, 2026

### **Section 4: Supplementary figures**

#### Figure S1

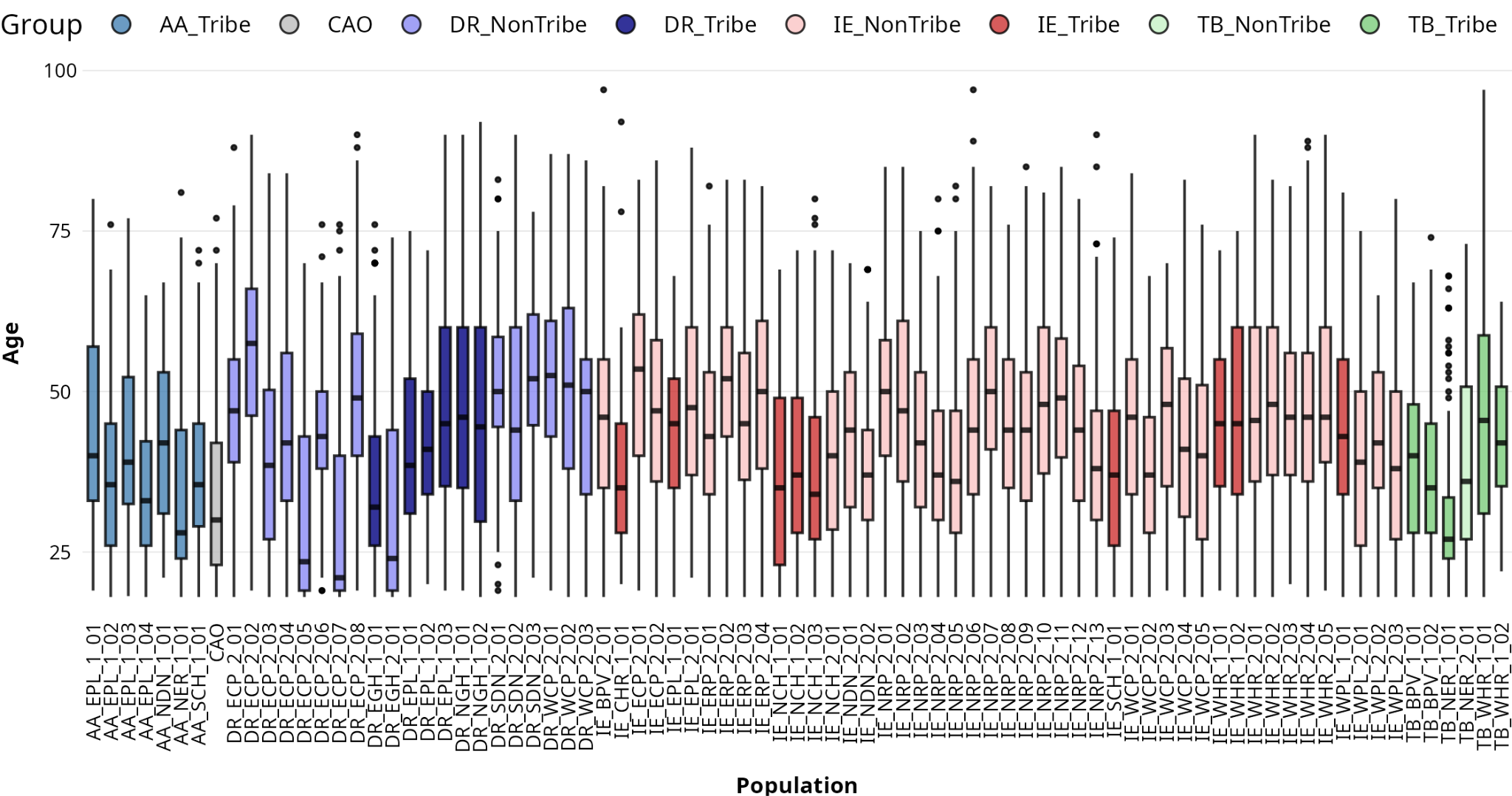

**Figure S1**. Age distributions of the 81 ethnolinguistic populations with phenotype data sampled for GI

#### Figure S2

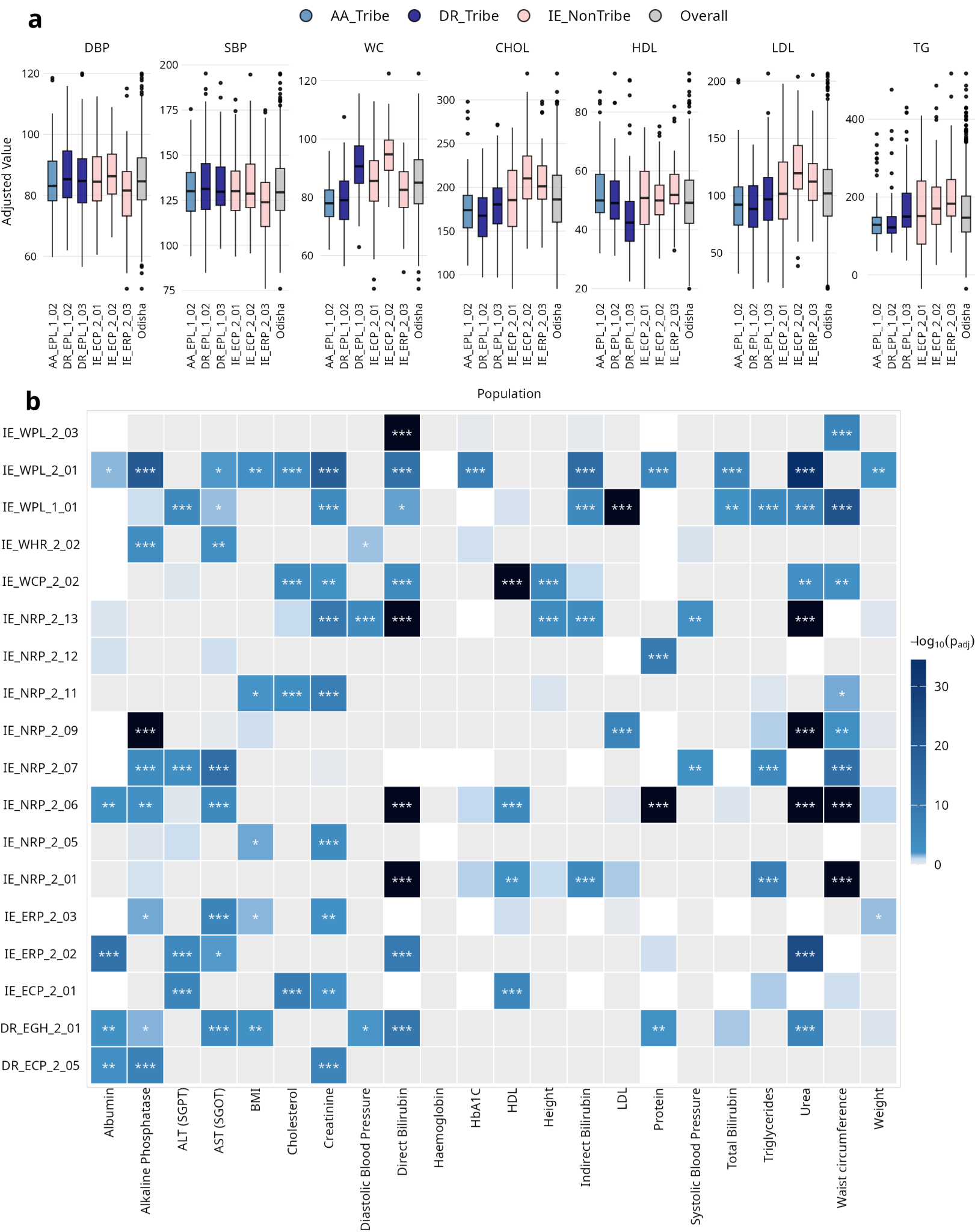

**Figure S2**. The degree of geographic phenotypic distribution is trait dependent. a: Variation in selected variables (HDL, LDL, Cholesterol, Triglycerides, Waist Circumference, Systolic and Diastolic BP) for the six populations surveyed from Odisha, subset for individuals not on medication. b: Heatmap showing the significance of phenotypic differences between two sampling states for each ethnolinguistic population (rows) and biomarker (columns). Colour intensity represents -log10(p-value) from a KS test; white cells indicate insufficient sample size or non-significant differences.

#### Figure S3

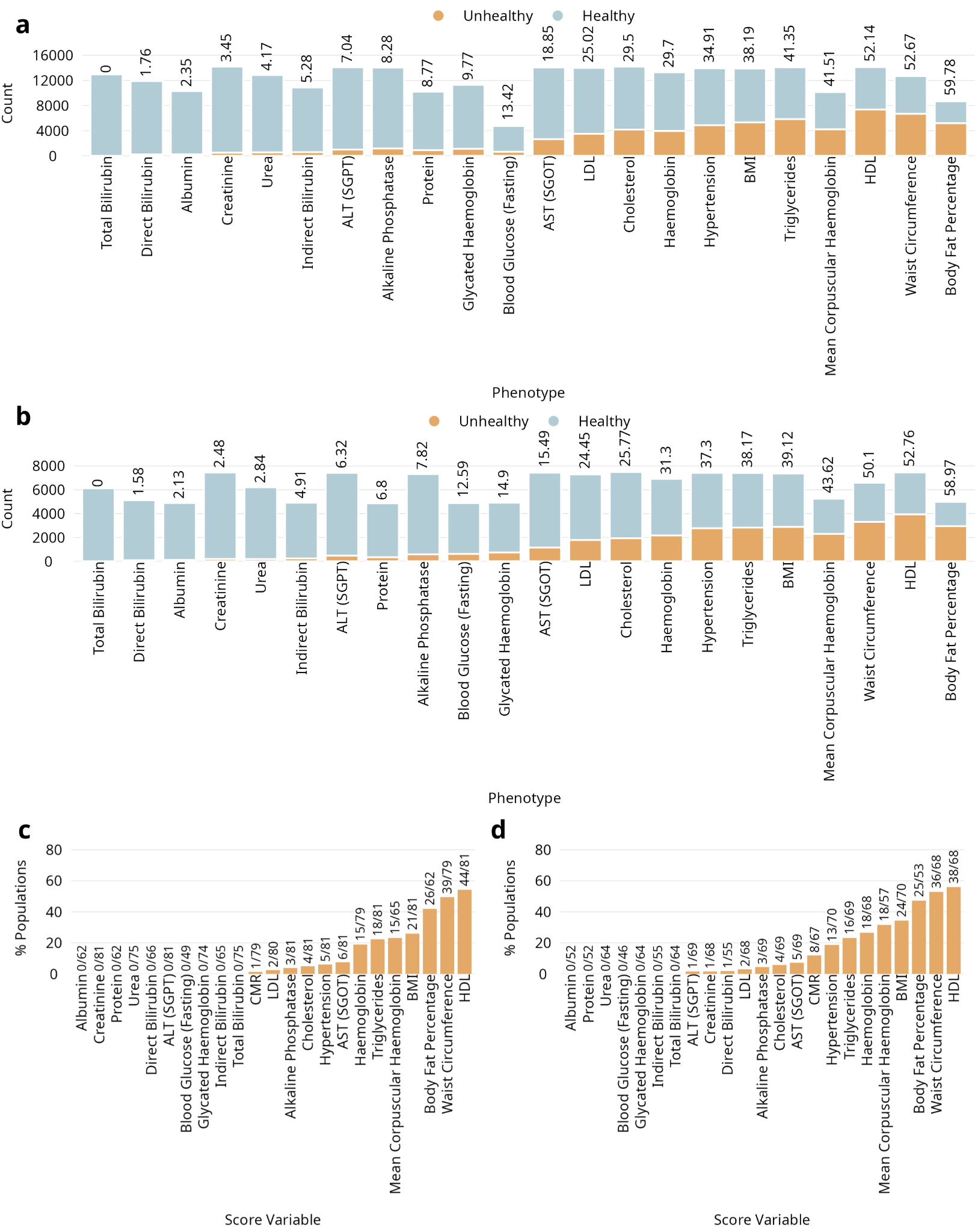

**Figure S3.** Prevalence of phenotypic abnormalities in the sampled populations. a: Counts of healthy and unhealthy samples for 22 phenotypes (both blood biochemistry and anthropometric, excluding blood cell counts), subset to the fraction of the individuals who were not on medication b: Counts of healthy and unhealthy samples for 22 phenotypes (both blood biochemistry and anthropometric, excluding blood cell counts), subset to the fraction of the individuals who were fasting c: Number of ethnicities that have 50% or more samples flagged as unhealthy for the 22 phenotypes in a, subset to the fraction of individuals not on medication. d: Number of ethnicities that have 50% or more samples flagged as unhealthy for the 22 phenotypes in b, subset to the fraction of individuals who were fasting.

#### Figure S4

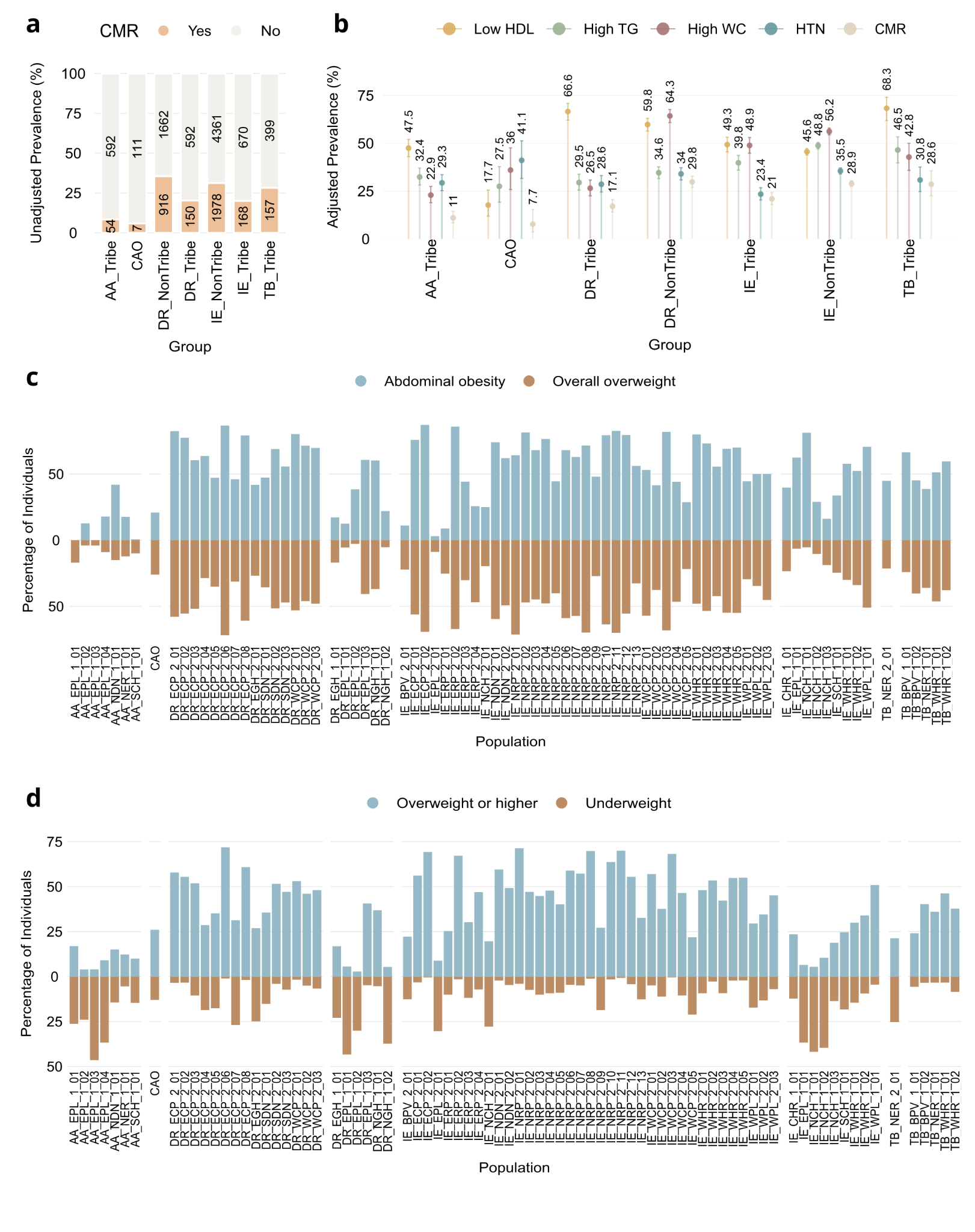

**Figure S4.** Metabolic risk profiles vary across populations and population groups a: Prevalence of clustered metabolic risk in each group for the individuals not on medication b: Age and gender-adjusted risk for each risk factor in the clustered metabolic risk symptoms for individuals not on medication c: Fraction of individuals overweight (BMI 25 or more) or higher vs abdominally obese in each sampled population d: Fraction of individuals overweight or higher (BMI 25 or more) vs underweight (BMI 18.5 or less) in each sampled population

#### Figure S5

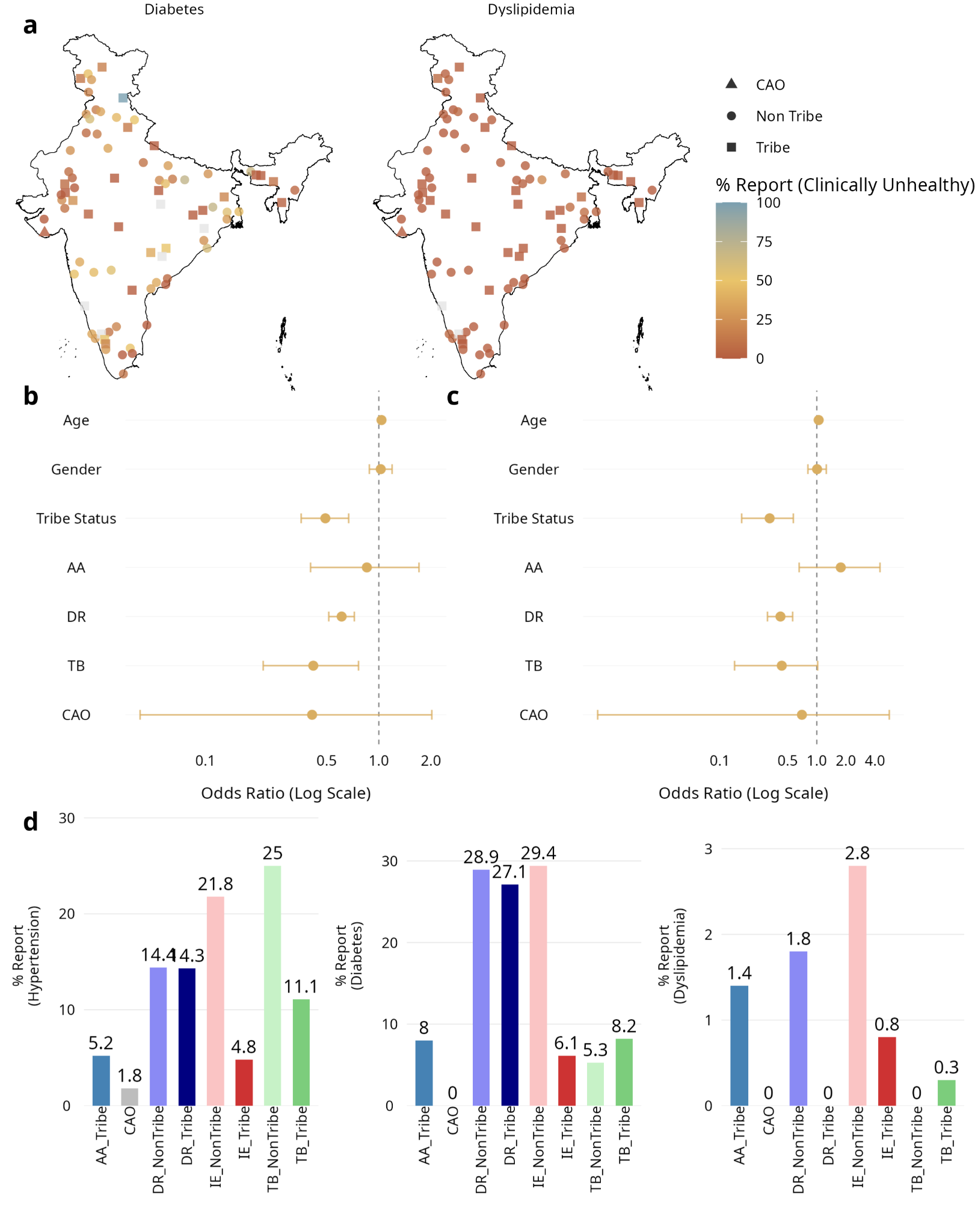

**Figure S5**. Variation in reporting fractions of diabetes, dyslipidemia, and hypertension across ancestry groups in GI a: Reporting fractions of diabetes and dyslipidemia for each sampled population in GI (grey dots indicate no information available). b: Odds ratio of awareness (reporting/medication) for diabetes, adjusting for age, gender, ancestry group and tribal status (reference set as IE, Non-Tribe, Female). c: Odds ratio of awareness (reporting/medication) for dyslipidemia, adjusting for age, gender, ancestry group and tribal status (reference set as IE, Non-Tribe, Female) d: unadjusted reporting fractions for hypertension, diabetes and dyslipidemia for the eight sampled ethnolinguistic groups

#### Figure S6

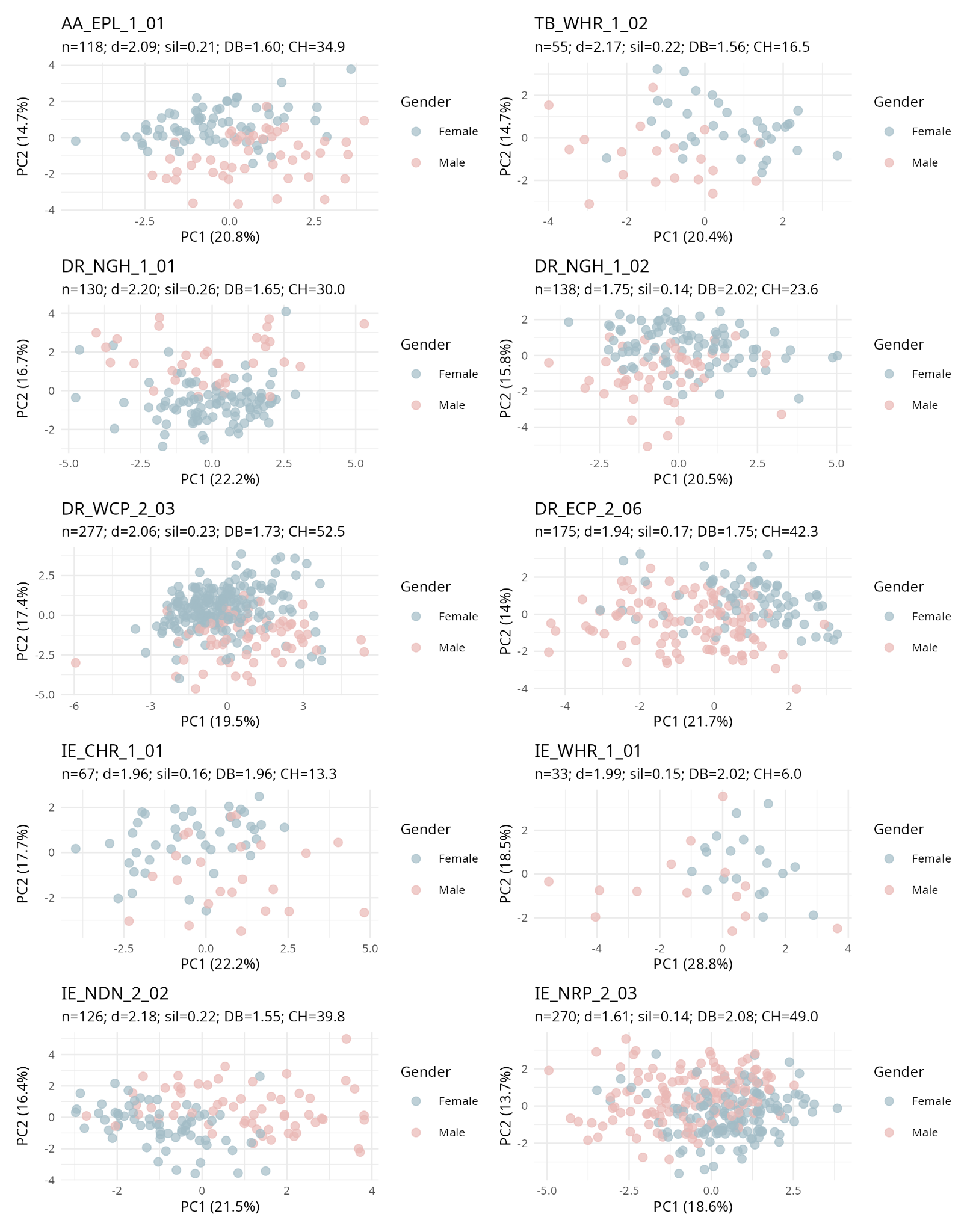

**Figure S6.** Extent of separation between genders via phenotype PCA, using representative ethnicities from each linguistic classification + tribe/non-tribe subgroup. (1 from Austro Asiatic Tribe, 1 from Tibeto Burman Tribe, 2 from Dravidian Tribe, 2 from Dravidian Non-Tribe, 2 from Indo European Tribe, 2 from Indo European Non-Tribe)

#### Figure S7

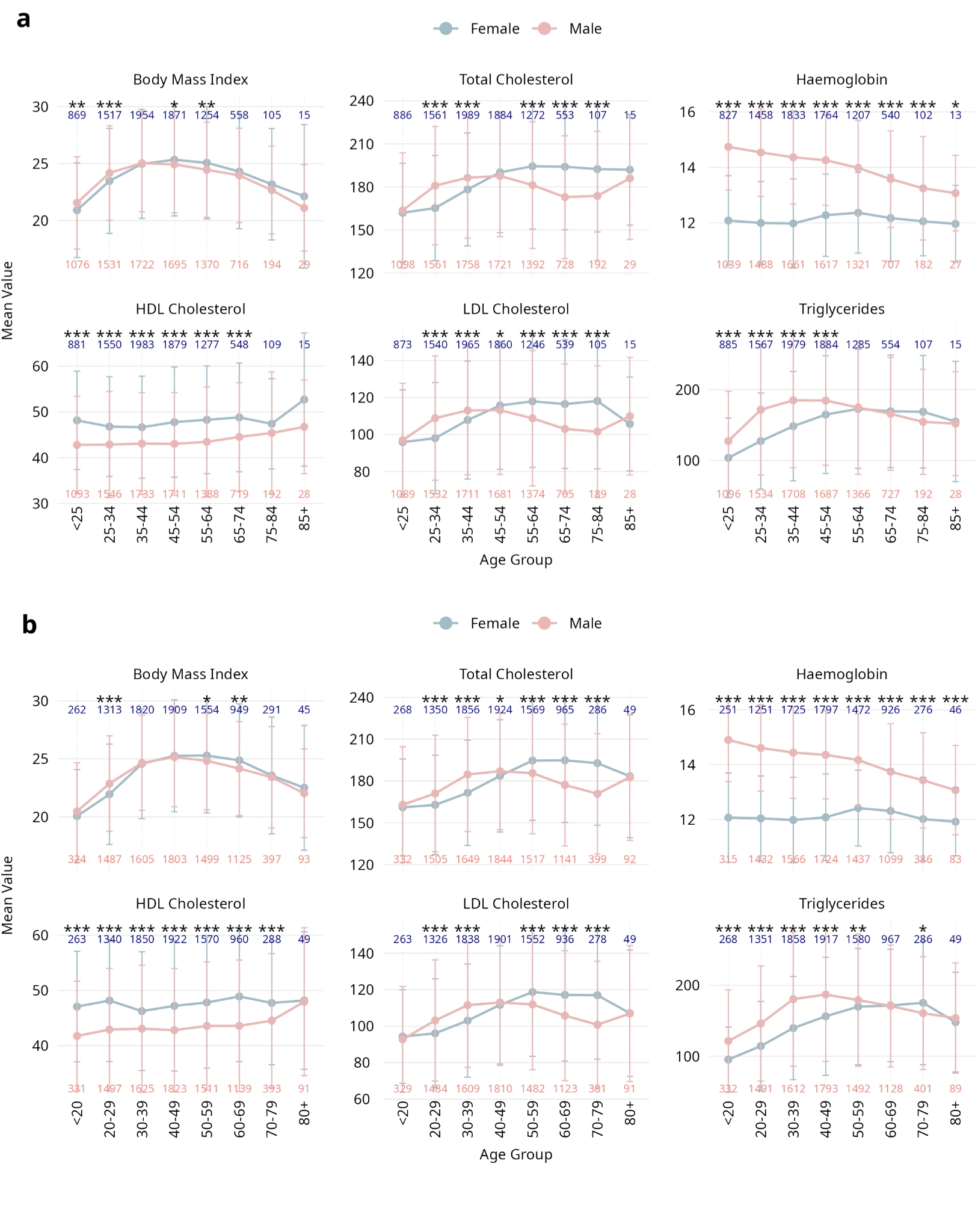

**Figure S7**. Life course trends of selected variables for males and females a: Trends remain significant for all individuals, irrespective of medication status, b: Trends remain robust across different choices of age bins. Significant differences in each age bin are determined by t-test and Benjamini-Hochberg correction for multiple testing (adjusted p<0.05 is significant. Adjusted p < 0.001 - ***, adjusted p < 0.01 - **, adjusted p < 0.05 - *)

#### Figure S8

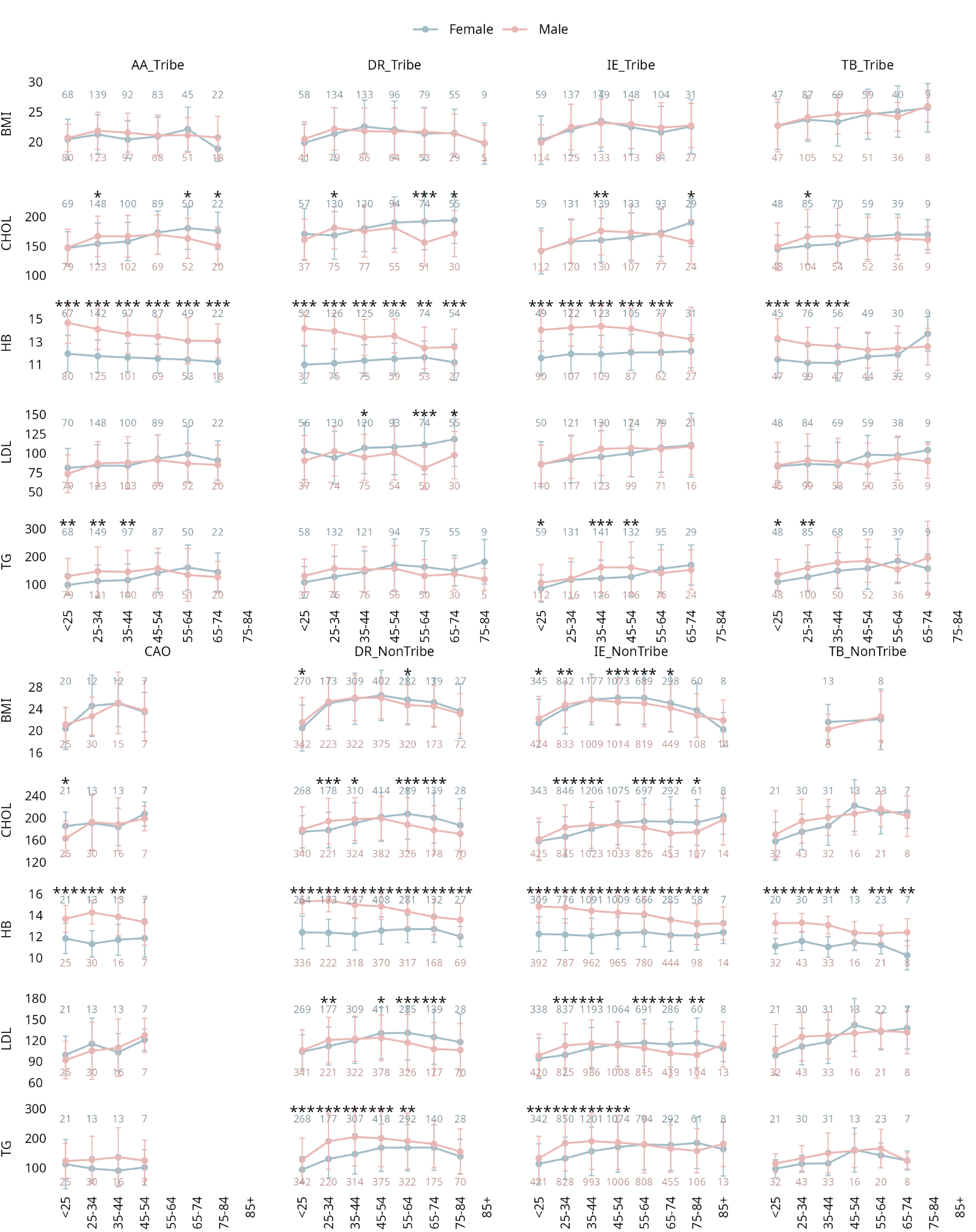

**Figure S8**. Life course trends of selected variables for males and females not on medication, for each ethnolinguistic group, for bins that have at least 5 male and female individuals. Significant differences in each age bin are determined by t-test and Benjamini-Hochberg correction for multiple testing (adjusted p<0.05 is significant. Adjusted p < 0.001 - ***, adjusted p < 0.01 - **, adjusted p < 0.05 - *)
